# The role of the SwissCovid digital proximity tracing app during the pandemic response: results for the Canton of Zurich

**DOI:** 10.1101/2021.02.01.21250972

**Authors:** Dominik Menges, Hélène Aschmann, André Moser, Christian L. Althaus, Viktor von Wyl

## Abstract

**Importance:** Digital proximity tracing (DPT) apps were released in several countries to help interrupt SARS-CoV-2 transmission chains in the population. However, the impact of DPT on pandemic mitigation still remains to be demonstrated.

**Objective:** To estimate key populations and performance indicators along the DPT app notification cascade in a clearly defined regional (Canton of Zurich, using all of Switzerland as a comparison) and temporal context (September/October 2020).

**Design:** Publicly available administrative and research data, including key DPT performance indicators, SARS-CoV-2 testing statistics, infoline call statistics, and observational study data, were compiled. A model of the DPT notification cascade was developed and key performance indicators for DPT processes were defined. Subpopulation sizes at each cascade step were estimated using data triangulation. Resulting estimates were systematically checked for internal consistency and consistency with other up- or downstream estimates in the cascade. Stochastic simulations were performed to explore robustness of results.

**Results:** For the Canton of Zurich, we estimate that 537 app users received a positive SARS-CoV-2 test in September 2020, of whom 324 received and entered a CovidCode. This triggered an app notification for an estimated 1374 proximity contacts and led to 722 infoline calls. In total, 170 callers received a quarantine recommendation, and 30 app users tested positive for SARS-CoV-2 after an app notification, reflecting a performance above the national level.

Based on this quantification, key performance indicators were evaluated. For September 2020, these analyses suggest that SwissCovid triggered quarantine recommendations in the equivalent of 5% of all exposed contacts placed in quarantine by manual contact tracing. Per 11 CovidCodes entered in the app, we estimate that almost 1 contact tested positive for SARS-CoV-2 upon app notification.

However, longitudinal indicator analyses demonstrate bottlenecks in the notification cascade, as capacity limits were reached due to large increases in SARS-CoV-2 incidence in October 2020.

**Conclusion:** Although requiring confirmation, our estimations on the number of notified proximity contacts receiving quarantine recommendations or testing positive after notification suggest relevant contributions to mitigating the pandemic. Increasing SwissCovid app uptake and improving notification cascade performance may further enhance its impact.

**Key points:** *Question:* What is the real-life impact of Digital proximity tracing (DPT) apps on interrupting SARS-CoV-2 transmission chains?

*Findings:* This data-informed simulation study found that, in the canton of Zurich, the number of app notified persons receiving a quarantine recommendation corresponds to the equivalent of up to 5% of all mandatory quarantined contacts identified by manual contact tracing. Furthermore, about 1 in 11 notification triggers led to SARS-CoV-2 testing of an exposed proximity contact who was consecutively tested positive.

*Meaning:* DPT apps exert a measurable impact that will further scale as more persons use the apps.

## Background

Switzerland was one of the first countries worldwide to release a digital proximity tracing (DPT) app (“SwissCovid”) on June 25, 2020 to complement manual contact tracing (MCT) in the ongoing SARS-CoV-2 pandemic.^1,2^ DPT apps in Germany, Italy, or Switzerland follow the “decentralized, privacy-preserving proximity tracing” blueprint (DP-3T, ^3^), which is supported by the Google Apple Exposure Notification Application Programming Interface (GAEN-API, ^4^). DP-3T-based apps send out Bluetooth Low Energy beacons that include regularly changing, anonymous identification numbers. Other DP-3T apps in the surroundings (“proximity contacts”) record and store these identification numbers. If an app user tests positive for SARS-CoV-2, this infected person can trigger exposure notifications to her/his proximity contacts through the DPT app in a privacy preserving manner. In the DP-3T architecture, anonymity of infected app users and proximity contacts is ensured by design because proximity encounter data are stored and evaluated locally on smartphones.^3^ The only centrally stored data are the anonymous identification numbers that are released by infected app users when releasing the exposure notification.

The exact steps necessary for warning proximity contacts through the DPT app (“notification cascade”) vary between countries. Figure 1 illustrates the notification cascade for Switzerland, one of the first countries to adopt DPT with the SwissCovid app. In Switzerland, app users who test positive for SARS-CoV-2 must request a notification code (CovidCode) from cantonal public health authorities. Entering the CovidCode into the app will lead to the upload of all identification numbers that were broadcast within the time window of infectiousness to a central server. DPT apps regularly download the list of all released identification numbers from the central server and compare it against the internally stored list of identifiers collected during recent proximity encounters. The matching procedure takes distance (as measured by Bluetooth signal strength attenuation) and duration of exposure into account. If the algorithm determines a significant exposure risk (i.e., <1.5m distance for 15 minutes or more within the window of infectiousness), the app will notify the user about the possible exposure.

**Figure 1:**
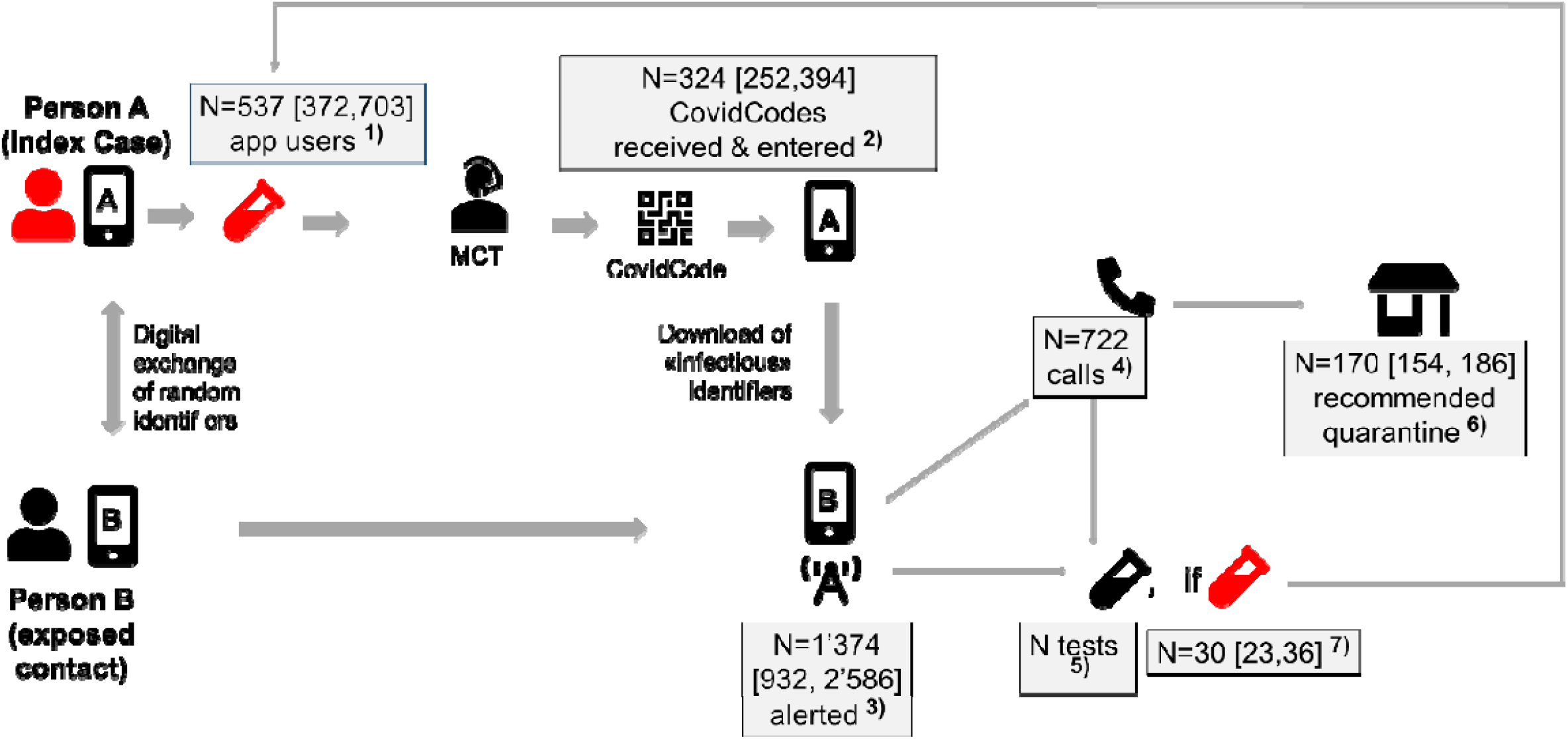
Conceptual model of the notification cascade. The notification cascade of the SwissCovid digital proximity tracing app consists of the following steps: **1)** An infected person A (index case) is tested positive for SARS-CoV-2 (red), is referred to manual contact tracing (MCT) and **2)** receives and uploads a CovidCode to warn other app users. Person B (exposed proximity contact) was in close proximity. **3)** This person receives the app notification, upon which he/she has several options: **4)** calling an infoline (recommended option), **5)** receiving a free test, and/or **6)** voluntary quarantine. **7)** Some persons will test positive after app notification, who will then be placed in isolation by manual contact tracing. The different populations at each step were calculated using data triangulation and stochastic simulation, as described in Table 2. The numbers reflect simulation results for the Canton of Zurich in September 2020 (first data column in Table 3). [Results in square brackets] reflect 2.5^th^ and 97.5^th^ percentiles of 50’000 simulations. Abbreviations: MCT, manual contact tracing.

In Switzerland, notified app users are eligible for a free SARS-CoV-2 test and are instructed to call an information hotline (“infoline”). The staff from the infoline will then determine whether a quarantine is warranted. However, the quarantine is not legally enforced. Thus, the effectiveness of the app in stopping transmission chains strongly depends on the actions taken by notified app users to prevent further possible transmissions, such as by entering self-quarantine.^2,5^

**Table 1:**
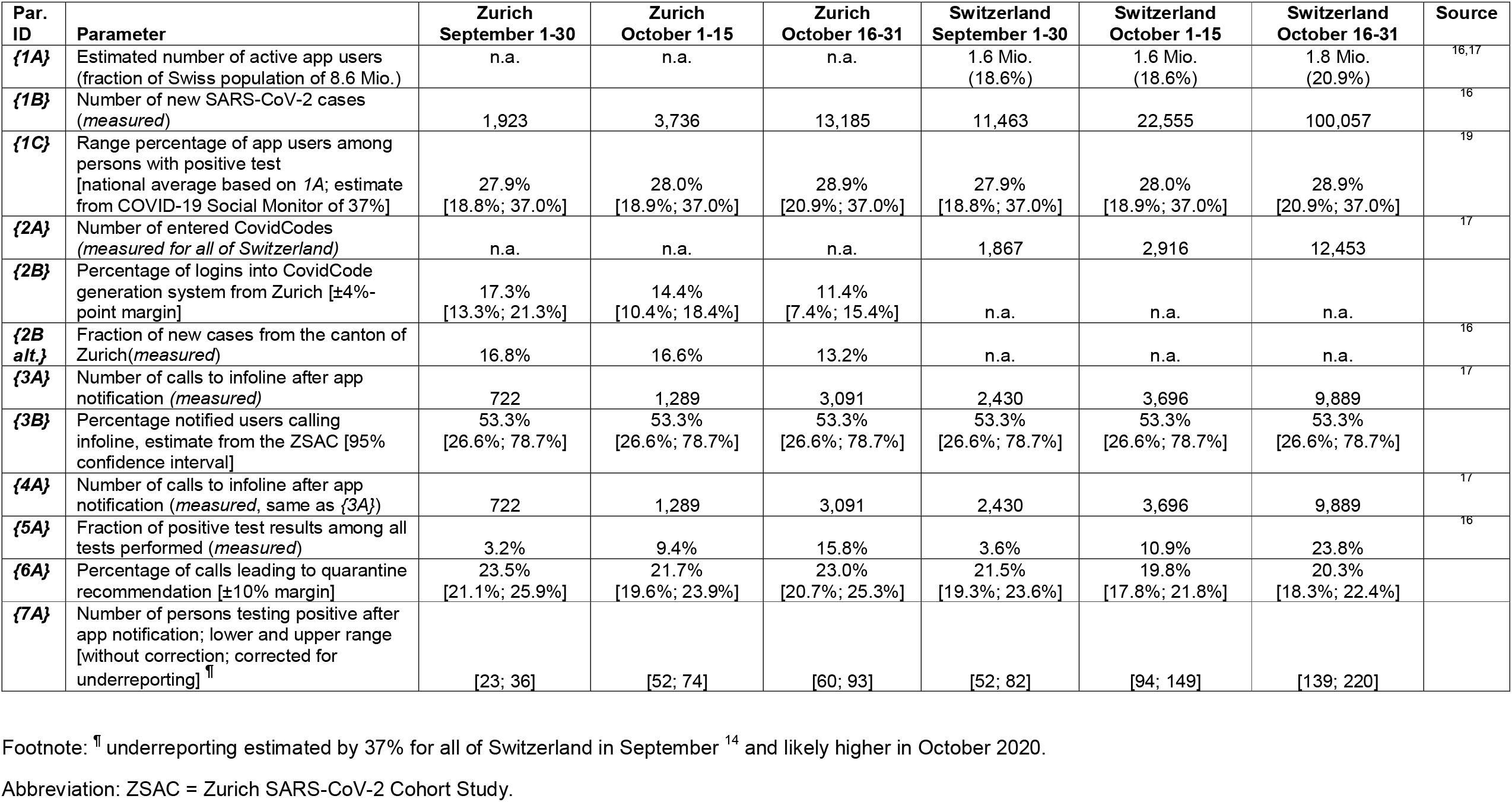
List of input parameters and uncertainty ranges. Parameters without uncertainty ranges reflect measured values.

| Par. ID | Parameter | Zurich<br>September 1-30 | Zurich<br>October 1-15 | Zurich<br>October 16-31 | Switzerland<br>September 1-30 | Switzerland<br>October 1-15 | Switzerland<br>October 16-31 | Source |
| --- | --- | --- | --- | --- | --- | --- | --- | --- |
| {1A} | Estimated number of active app users (fraction of Swiss population of 8.6 Mio.) | n.a. | n.a. | n.a. | 1.6 Mio.<br>(18.6%) | 1.6 Mio.<br>(18.6%) | 1.8 Mio.<br>(20.9%) | <sup>16,17</sup> |
| {1B} | Number of new SARS-CoV-2 cases (*measured*) | 1,923 | 3,736 | 13,185 | 11,463 | 22,555 | 100,057 | <sup>16</sup> |
| {1C} | Range percentage of app users among persons with positive test [national average based on 1A; estimate from COVID-19 Social Monitor of 37%] | 27.9%<br>[18.8%; 37.0%] | 28.0%<br>[18.9%; 37.0%] | 28.9%<br>[20.9%; 37.0%] | 27.9%<br>[18.8%; 37.0%] | 28.0%<br>[18.9%; 37.0%] | 28.9%<br>[20.9%; 37.0%] | <sup>19</sup> |
| {2A} | Number of entered CovidCodes (*measured for all of Switzerland*) | n.a. | n.a. | n.a. | 1,867 | 2,916 | 12,453 | <sup>17</sup> |
| {2B} | Percentage of logins into CovidCode generation system from Zurich [±4%-point margin] | 17.3%<br>[13.3%; 21.3%] | 14.4%<br>[10.4%; 18.4%] | 11.4%<br>[7.4%; 15.4%] | n.a. | n.a. | n.a. |  |
| {2B alt.} | Fraction of new cases from the canton of Zurich (*measured*) | 16.8% | 16.6% | 13.2% | n.a. | n.a. | n.a. | <sup>16</sup> |
| {3A} | Number of calls to infoline after app notification (*measured*) | 722 | 1,289 | 3,091 | 2,430 | 3,696 | 9,889 | <sup>17</sup> |
| {3B} | Percentage notified users calling infoline, estimate from the ZSAC [95% confidence interval] | 53.3%<br>[26.6%; 78.7%] | 53.3%<br>[26.6%; 78.7%] | 53.3%<br>[26.6%; 78.7%] | 53.3%<br>[26.6%; 78.7%] | 53.3%<br>[26.6%; 78.7%] | 53.3%<br>[26.6%; 78.7%] |  |
| {4A} | Number of calls to infoline after app notification (*measured*, same as {3A}) | 722 | 1,289 | 3,091 | 2,430 | 3,696 | 9,889 | <sup>17</sup> |
| {5A} | Fraction of positive test results among all tests performed (*measured*) | 3.2% | 9.4% | 15.8% | 3.6% | 10.9% | 23.8% | <sup>16</sup> |
| {6A} | Percentage of calls leading to quarantine recommendation [±10% margin] | 23.5%<br>[21.1%; 25.9%] | 21.7%<br>[19.6%; 23.9%] | 23.0%<br>[20.7%; 25.3%] | 21.5%<br>[19.3%; 23.6%] | 19.8%<br>[17.8%; 21.8%] | 20.3%<br>[18.3%; 22.4%] |  |
| {7A} | Number of persons testing positive after app notification; lower and upper range [without correction; corrected for underreporting] <sup>¶</sup> | [23; 36] | [52; 74] | [60; 93] | [52; 82] | [94; 149] | [139; 220] |  |

Footnote: <sup>¶</sup> underreporting estimated by 37% for all of Switzerland in September <sup>14</sup> and likely higher in October 2020.
Abbreviation: ZSAC = Zurich SARS-CoV-2 Cohort Study.

**Table 2:** Calculation process.

| Notification Cascade Step | Sub-population to be estimated | Estimation process<br>(numbers in *{brackets}* refer to parameter IDs in Table 1) |
| --- | --- | --- |
| 1) | Number of app users who test positive for SARS-CoV-2 | Number of positive cases *{1B}*<br>*multiplied by*<br>Fraction of app users in Swiss population *{1A}* |
| 2) | Number of CovidCodes entered in app | Number of total CovidCodes entered in Switzerland *{2A}*<br>*multiplied by*<br>Percentage of logins into CovidCode generation software by Zurich as a proxy for code generation *{2B}*<br>(alternative: Fraction of positive cases from Canton of Zurich *{2B alt.}*) |
| 3) | Number of persons notified by app | Number of infoline calls *{3A}*<br>*divided by*<br>Percentage of persons calling the infoline among all persons notified by the app *{3B}* |
| 4) | Number of infoline calls related to app exposure notifications | Number of infoline calls *{3A}* |
| 5) | Number of tests performed upon app notification | Number of positive tests upon app notification *{7A}*<br>*divided by*<br>Percentage of positive tests among all tests performed *{5A}* <sup>¶</sup> |
| 6) | Number of infoline calls leading to quarantine recommendation | Number of infoline calls *{3A}*<br>*multiplied by*<br>Percentage of calls leading to quarantine recommendation *{6A}* |
| 7) | Number of positive tests after app notification | Number of positive test results where test was performed after app notification *{7A}* |

Footnote: <sup>¶</sup> The overall test positivity rate is likely an underestimation of the true positivity for persons with digital proximity contact exposure.

**Table 3:** Quantification of the SwissCovid app notification cascade and estimation of monitoring parameters.

| Step | Outcomes | Zurich,<br>September 1-30<br>Median<br>[2.5<sup>th</sup>, 97.5<sup>th</sup>]<br>percentiles | Zurich,<br>October 1-15<br>Median<br>[2.5<sup>th</sup>, 97.5<sup>th</sup>]<br>percentiles | Zurich,<br>October 16-31<br>Median<br>[2.5<sup>th</sup>, 97.5<sup>th</sup>]<br>percentiles | Switzerland<br>September 1-30<br>Median<br>[2.5<sup>th</sup>, 97.5<sup>th</sup>]<br>percentiles | Switzerland<br>October 1-15<br>Median<br>[2.5<sup>th</sup>, 97.5<sup>th</sup>]<br>percentiles | Switzerland<br>October 16-31<br>Median<br>[2.5<sup>th</sup>, 97.5<sup>th</sup>]<br>percentiles |
| --- | --- | --- | --- | --- | --- | --- | --- |
| <u>Steps in notification cascade</u> |  |  |  |  |  |  |  |
| 1) | Number of app users among persons with positive test | 537<br>[372; 703] | 1,042<br>[722; 1,365] | 3,818<br>[2,809; 4,822] | 3,205<br>[2,209; 4,191] | 6,296<br>[4,365; 8,249] | 28,931<br>[21,302; 36,602] |
| 2) | Number of entered CovidCodes | 324<br>[252; 394] | 422<br>[310; 532] | 1,426<br>[951; 1,896] | 1,867 | 2,916 | 12,453 |
| 3) | Number of notified app contacts | 1,374<br>[932; 2,586] | 2,457<br>[1,665; 4,619] | 5,886<br>[3,996; 11,062] | 4,633<br>[3,138; 8,707] | 7,020<br>[4,773; 13,244] | 18,707.5<br>[12,772; 35,445] |
| 4) | Number of calls to infoline related to app notification | 722 | 1,289 | 3,091 | 2,430 | 3,696 | 9,889 |
| 5) | Number of tests due to an app notification | 939<br>[720; 1,127] | 626<br>[509; 743] | 977<br>[406; 1,549] | 3,733<br>[1,560; 5,906] | 2,378<br>[933; 3,814] | 4,255<br>[770; 7,764] |
| 6) | Number of calls to infoline leading to quarantine recommendation | 170<br>[154; 186] | 280<br>[253; 307] | 711<br>[643; 778] | 522<br>[472; 572] | 733<br>[663; 803] | 2,015<br>[1824; 2207] |
| 7) | Number of persons testing positive for SARS-CoV-2 upon app notification | 30<br>[23; 36] | 67<br>[53; 82] | 78<br>[61; 94] | 67<br>[53; 82] | 121<br>[95; 148] | 180<br>[141; 218] |
| <u>Other derived performance indicators</u> |  |  |  |  |  |  |  |
|  | Fraction of entered CovidCodes per SARS-CoV-2 positive app user | 60.3%<br>[39.5%; 96.5%] | 40.4%<br>[25.3%; 66.0%] | 37.3%<br>[22.2%; 60.5%] | 58.3%<br>[44.5%; 84.5%] | 46.3%<br>[35.3%; 66.8%] | 43.0%<br>[34.0%; 58.5%] |
|  | Ratio of notified app users per entered CovidCode | 4.3<br>[2.6; 8.7] | 6.0<br>[3.4; 12.4] | 4.3<br>[2.3; 9.3] | 2.5<br>[1.7; 4.7] | 2.4<br>[1.6; 4.5] | 1.5<br>[1.0; 2.8] |
|  | Ratio of entered CovidCodes per app user testing positive upon app notification | 10.9<br>[7.6; 15.6] | 6.2<br>[4.2; 9.3] | 18.4<br>[11.2; 28.3] | 27.9<br>[22.8; 35.2] | 24.1<br>[19.7; 30.7] | 69.2<br>[57.1; 88.3] |
|  | Ratio of the number of infoline callers receiving a quarantine recommendation over the number of all quarantined persons. | 5.1%<br>[4.6%; 5.6%] | 3.5%<br>[3.1%; 3.8%] | 3.1%<br>[2.8%; 3.4%] | 2.0%<br>[1.8%; 2.2%] | 1.7%<br>[1.6%; 1.9%] | 2.0%<br>[1.8%; 2.2%] |
|  | Ratio of infoline calls per entered CovidCode (all of Switzerland) | n.a. | n.a. | n.a. | 1.54 | 1.60 | 1.01 |
|  | Ratio of persons tested positive upon app notification over all SARS-CoV-2 positive cases | 1.6%<br>[1.2%; 1.9%] | 1.8%<br>[1.4%; 2.2%] | 0.6%<br>[0.5%; 0.7%] | 0.6%<br>[0.5%; 0.7%] | 0.5%<br>[0.4%; 0.7%] | 0.2%<br>[0.1%; 0.2%] |

In principle, DPT has several advantages over MCT. First, SARS-CoV-2 positive app users do not have to rely on memory to recall proximity contacts. Second, once the positive test result is known to the user, notification happens almost in real-time and simultaneously for all proximity contacts, without requiring human resources.^5^ Ultimately, DPT may thus be faster and have a wider reach than the more labor-intensive MCT. Along those lines, several modelling studies suggest that DPT apps have the potential to slow or even stop pandemic transmission ^2,6-8^, provided that a substantial fraction of the population uses the app and swiftly complies with recommended actions (e.g. going into quarantine upon app notification).

However, the reality is more complex. ^9^ In Switzerland, the notification cascade involves user interactions that are explicitly voluntary and where user incentives may play an important role.^10,11^ Notified app users are eligible for a free SARS-CoV-2 test. But contrary to quarantine mandated by the public health authorities as part of MCT, the (voluntarily) quarantine following an app notification is not eligible for salary compensation in Switzerland (which is currently being reconsidered).^12^ Emerging first evidence also suggests that DPT effectiveness may be affected by bottlenecks in the notification cascade and incomplete compliance with recommended actions.^13^ Furthermore, app usage seems to plateau even in countries with comparatively high app adoption. If measured by DPT app downloads, population coverage is estimated at around 30% for Switzerland and Germany (and even lower if only the number of active apps are counted, with 21% for Switzerland). However, some modelling studies argue that even a coverage of 20-30% may suffice to produce some effects if combined with other pandemic mitigation measures (particularly MCT).^8^

Meanwhile, early analyses based on routine monitoring data demonstrate that DPT in principle can work as intended. A study by Salathé and colleagues shows that, between July and September 2020, 45 persons were tested positive after an app notification in Switzerland.^14^ While this can be seen as proof-of-principle that the technical aspects DPT work, a more detailed quantification and contextualization of its effects is warranted.^14^ Early population-level data are provided by study from the Isle of Wright, which analyzed the impact of the implementation of its “Test-and-Trace” program in May 2020, which included a DPT app.^15^ While this study shows that the combined measures clearly had a lowering impact on SARS-CoV-2 incidence, the contribution of DPT was difficult to evaluate.

Therefore, empirical studies of the effectiveness of DPT apps and a quantification of their population-level impacts are urgently needed. Against this background, our analysis has two aims. First, by combining data from different sources, we aimed to describe the different steps in the notification cascade and quantify the respective population in each step for a clearly defined regional and temporal context. Second, building on these results we identified and estimated possible indicators to advance our understanding of the impact of DPT relative to MCT. The analysis was conducted during a period of a marked rise in SARS-CoV-2 incidence, which brought MCT, testing laboratories and health facilities in some areas to capacity limits. Therefore, our study also enabled a closer analysis of how possible bottlenecks in the health systems may affect the notification cascade.

## Methods

### Setting

We analyzed data collected between September 1 and October 31, 2020. As illustrated by Figure 2, daily SARS-CoV-2 incidence in Switzerland was relatively stable at 300 to 600 new cases in September (blue line), followed by a steep increase in October to 10,000 casesand a decrease in November (not shown). ^16^ Figure 2 further shows the number of entered CovidCodes (red line) and infoline calls (green line), which largely followed the trajectory of SARS-CoV-2 incidence. However, CovidCode and infoline call curves crossed in October, which was caused by a temporary overload at the infoline call center due to high call volumes. There were also delays in issuing CovidCodes for positive tested app users, which has been documented by an increasing time from symptom onset to CovidCode upload. ^17^ Due to these capacity issues mentioned above, the analysis for the month of October was conducted for two separate time periods (October 1 to 15 and October 16 to 31, 2020).

**Figure 2:**
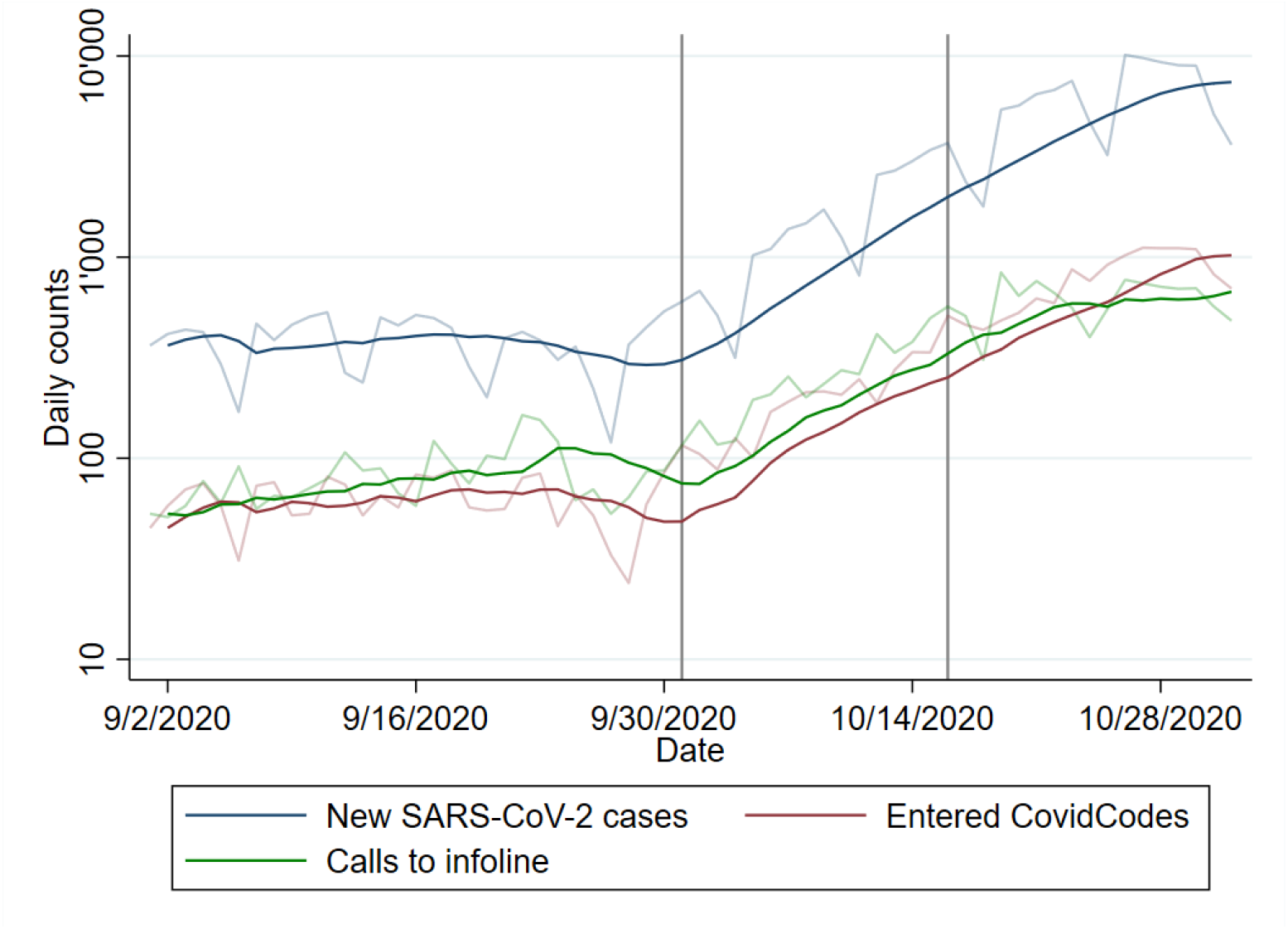
Evolution of key statistics in Switzerland for the months of September and October 2020. Transparent lines are numbers from daily reporting, solid lines reflect 7-day moving averages. Vertical grey lines mark cut-off dates for the different time strata utilized in the analysis.

The primary study focus was the notification cascade in the Canton of Zurich with 1.54 Million inhabitants (compared to 8.6 Million inhabitants in Switzerland overall). The Canton of Zurich was selected due to the availability of several, relevant data sources from public health administration and research (see section “Data Sources”). ^18,19^

### Data sources

Key epidemiological data on app usage and indicators of app effectiveness were extracted from various sources and are summarized in Table 1 for both the Canton of Zurich and Switzerland overall (with numbers in {curly brackets} corresponding to specific parameters in Table 1):

1. <u>Federal Office of Public Health (FOPH)</u>. Data included the number of positive SARS-CoV-2 tests by age group and canton ***{1B}***. ^16^ The FOPH collects information on testing reasons for positive SARS-CoV-2 tests, including tests that noted a SwissCovid app alert as a reason for testing ***{7A}***. These data were provided directly to the main author.
2. <u>Federal Office of Statistics (FOS)</u>. Publicly available data included the number of active SwissCovid app users and the number of entered notification codes (CovidCodes) ^17^.
3. <u>Cantonal Health Directorate Zurich (GDK-ZH)</u>. The GDK-ZH compiles data on positive cases and operates MCT. Obtained data from this source include the number of positive cases in the Canton of Zurich ***{1B}***, the ratio of quarantined and isolated persons, the number of tests performed in Zurich and test positivity ***{5A}***.^20,21^
4. <u>Medgate</u> is a private company operating the national infoline, which users notified by the app are encouraged to call. It maintains statistics on number of infoline calls pertaining to app notifications ***{3A}*** and the percentage of infoline calls leading to quarantine recommendation ***{6A}***. While the number of calls is public ^17^, other information were provided directly to the main author.
5. <u>COVID-19 Social Monitor</u> is a longitudinal panel survey on COVID-19 in Switzerland, that includes between 1,500 to 1,700 respondents per wave.^22^ The survey was started in March 2020, and 11 waves were completed by November 2020. It provided aggregated statistics on app usage ***{1A}*** and the percentage of app users calling the infoline after notification ***{3B}***. The main author had direct access to the data.
6. <u>Zurich SARS-CoV-2 Cohort (ZSAC) Study</u> is a longitudinal cohort study embedded in contact tracing of the Canton of Zurich ^23^ and provided aggregated data on the percentage of notified app users who called infoline ***{3B}*** and the percentage of index cases that were tested positive after an app notification ***{7A}***. The main author received the data directly.

### Calculation Method

This study employed data triangulation ^24^, combined with stochastic simulations to explore the robustness of estimates. The triangulation process followed the framework of Kaufmann et al. ^25^ Key outcome measures were defined as described in Table 2, a list of possible administrative and research data sources was compiled, data sources were screened for relevant information, and suitable parameter data was extracted.

Next, a schematic model for the notification cascade was defined, outlining the flow of users and information (Figure 1; analogous to the model presented elsewhere ^14^).

For study aim 1, the respective population size was calculated for each step in the notification cascade, based on measured data and estimated parameters (Table 2). In addition, the following complementary performance indicators were defined for study aim 2:

- Fraction of entered CovidCodes per SARS-CoV-2 positive app user.
- Ratio of notified app users per entered CovidCode.
- Ratio of entered CovidCodes per app user testing positive after app notification.
- Ratio of the number of infoline callers receiving a quarantine recommendation over the number of all quarantined persons.
- Ratio of infoline calls per entered CovidCode.
- Ratio of persons tested positive upon app notification over all SARS-CoV-2 positive cases.

The triangulation was performed separately for the three different time periods (September, first and second half of October) and geographic entities (Canton of Zurich and Switzerland overall). It centered around two key indicators, for which measured data were available: the number of infoline calls from the Canton of Zurich (cascade step 4, parameter ***{4A}***) and the number of positive tests following an app notification in Zurich (step 7, parameter ***{7A}***; note that the measure is likely an underestimation ^14^). The triangulation process consisted of a size estimation of the remaining five subpopulations in the notification cascade). For estimated parameters (Table 1), uncertainty ranges were either based on 95% confidence intervals if parameters were derived from individual-level data, or by adding a 4%-10% margin around the best available data point if derived from aggregated data (with margin adjustment during the triangulation process described below).

Based on the parameters and uncertainty ranges, a stochastic simulation with 50,000 repetitions was performed, whereby parameter values were drawn at random from uniform parameter distributions defined by pre-specified parameter limits (Table 1). The estimated sizes of the five subpopulations in the cascade and the performance indicators were summarized as medians [2.5^th^ and 97.5^th^ percentiles; including 95% of all estimates] of parameter estimates across all simulations. The simulation results were systematically checked for consistency based on the following criteria (in descending order of priority):

1. Is the estimate consistent with independently collected, analogous data?
2. Is the estimate contradicting other up- or downstream estimates?
3. Are the estimates largely consistent with qualitative information if available?

In case of inconsistencies the simulation was iteratively refined by adjusting the uncertainty ranges. Calculations were performed in R version 3.6.2 (www.r-project.org).

## Results

First, the population sizes at the different notification cascade steps were estimated. Figure 1 shows subpopulation sizes of the individual cascade steps for the Canton of Zurich in September 2020. Based on our simulations, 537 of 1923 persons tested positive for SARS-CoV-2 were app users. Out of those, 324 (60.3%) received and entered a CovidCode, which triggered exposure notifications in 1,438 exposed proximity contacts. These notifications led to 722 calls to the infoline, out of which 170 (23.5%) led to a quarantine recommendation. Furthermore, 30 app users tested positive for SARS-CoV-2 after receiving the app notification.

Analogous calculations for the month of October and all of Switzerland are shown in Table 3 (upper half). As demonstrated in Figure 2 and Table 1 (second row), the number of SARS-CoV-2 positive cases in each period rose substantially in the first and second half of October in the Canton of Zurich (from 1,923 cases to 3,736 and 13,185 cases, respectively) and all of Switzerland (from 11,463 to 22,555 and 100,057 cases, respectively). The absolute sizes of notification cascade subpopulations also grew markedly between September and October. The estimated number of entered CovidCodes in Zurich increased from 324 to 1,426 between September and the second half of October, the number of infoline calls from 722 to 3,091, and the number of quarantine recommendations from 170 to 711.

We additionally conducted analyses of complementary performance indicators aiming to compare and evaluate the changes in subpopulation sizes with respect to the dynamics of the pandemic. These analyses, shown in the lower half of Table 3, suggest that the number of entered CovidCodes did not grow proportionally to the number of new SARS-CoV-2 cases. In the Canton of Zurich, the fraction of entered CovidCodes per SARS-CoV-2 positive app user changed from 60.3% in September to 40.4% and 37.3% in the first and second half of October. Corresponding numbers for all of Switzerland were 58.3%, 46.4% and 43.0%, respectively.

Longitudinal indicator changes were also observed for the ratio of the number of infoline callers receiving a quarantine recommendation over the number of all quarantined persons, which declined from 5.1% to 3.1% in Zurich (while the nationwide ratio remained stable at 2%). Similarly, the ratio of app users who tested positive after an app notification over all persons newly tested positive changed from 1.6% to 0.6% in Zurich and from 0.6% to 0.2% in Switzerland. Similar dynamics were mirrored by the ratio of entered CovidCodes per app user who tested positive upon app notification. In Zurich, 10.9 entered CovidCodes led to 1 positive test post-notification in September. This ratio decreased to 6.2 in the first half of October, before increasing again to 18.4. The nationwide ratio showed a similar dynamic but were 3 to 4 times higher (27.9 entered CovidCodes per detected case in September, 69.2 in the second half of October).

## Discussion

Several months into the release of the first DPT apps our understanding of the effectiveness and the impact of DPT on pandemic mitigation is still limited. Based on the premise that DPT is a complex public health intervention with a notification cascade consisting of several steps, the present study, provides a first estimation of the contribution of the SwissCovid DPT app to mitigating SARS-CoV-2 transmission in the Canton of Zurich.

During a period of relatively stable SARS-CoV-2 incidence in September 2020, we found that app notifications may have contributed to actions to prevent further viral transmission in 30 infected persons who were tested for SARS-CoV-2 upon app notification. Furthermore, we estimated that SwissCovid notifications led to self-quarantine recommendations in 170 persons. Overall, this estimate implies that the app could have led, at most, to a 5% increase of persons entering quarantine. Meanwhile, the effort to identify these persons likely was less labor- and resource-intensive than contact identification through MCT. Of note, the data from Zurich suggest an above-average performance of the app notification cascade in the Canton when compared to the nationwide DPT indicators. Therefore, our data provides solid first evidence that the DPT technology may be an effective complementary measure to identify infected cases and mitigate the spread of SARS-CoV-2 under real-world conditions.

However, the month of September was followed by 20- to 30-fold increase in SARS-CoV-2 incidence by the end of October, which led to temporary capacity issues for MCT, infoline call centers, and the generation of CovidCode. These bottlenecks were also reflected by a deterioration of most outcome indicators monitored by our study. Nevertheless, the rising SARS-CoV-2 incidence also had positive consequences for the adaptation and process efficiency of the DPT technology. From mid-August to mid-October 2020, the number of active app users in Switzerland had plateaued at 1.6 million. Renewed public appeals by health authorities for using the app resulted in 200,000 additional users by the end of October ^17^. Moreover, important bottlenecks in the notification cascade, most especially the capacity of the infoline and the process for issuing CovidCodes, were identified as a result of the increasing incidence. This led to the swift implementation of specific measures to improve DPT processes, such as technical solutions to handle higher call volumes and to automatize CovidCode generation, as well as introducing the possibility for any physician and infoline to issue CovidCodes if the cantonal health authorities are unable to respond in a timely manner.

Furthermore, Cantonal public health authorities scaled up MCT by hiring more personnel and automated different steps in contact tracing through an online form for index cases and close contacts. While the effects of these changes became fully visible after the conclusion of this analysis, some indicators from official statistics already suggest a recovery in the SwissCovid notification cascade performance. By November, the nationwide average time from symptom onset to CovidCode entry decreased to around 4 days after an average of 5 days in late October (not shown, ^17^). Furthermore, due to the introduction of rapid antigen testing at easily accessible sites (e.g., including 40 pharmacies in the canton of Zurich) this time interval is expected to drop further. These examples underscore the need for a continuous, comprehensive evaluation of the full DPT notification cascade on one side, but also for a fast adjustment of processes and increase in resources in case of capacity bottlenecks. In addition, increased automation of the different steps in the notification cascade may help to achieve better performance. For example, DPT systems in use in Germany and Belgium have automated the registration and communication of SARS-CoV-2 test results or the upload of temporary exposure keys in infected persons, while still obtaining explicit app user consent for these steps.^26,27^ Similar optimizations are underway in the Canton of Zurich.

To our knowledge, our study is the first systematic attempt to longitudinally quantify and evaluate the performance of the DPT app notification cascade. Our study is based on the premise that DPT apps are a complex intervention that depends on actions of app users and other actors in the health system. A media analysis from Switzerland previously reported on implementation challenges for DPT, including delays in the provision of CovidCodes to positively tested app users. ^13^ Our study confirms these observations and presents indicators for a more targeted monitoring of specific procedures along the notification cascade that pose potential bottlenecks.

Of further note, the calculation methods and indicators employed in this study for monitoring DPT performance do not require individual-level data and are thus fully compatible with the privacy-preserving design and philosophy of DP-3T. However, this is also a limitation. For example, the large increase in SARS-CoV-2 incidence in the second half of October also affected the data collection procedures for monitoring statistics. For example, the reporting of the reasons for testing (such as a SwissCovid app notification), which primarily relies on physicians diagnosing the infection, was already incomplete in September but nearly stopped towards the end of October 2020. This underreporting also may have led to the underestimation of some indicators in our analysis (e.g. the number of persons testing positive after app notification). However, we partly accounted for this issue through the performance of stochastic analysis and the presentation of uncertainty ranges.

Furthermore, some of the parameter estimates utilized in our calculations were derived from studies with still limited sample sizes and follow-up (e.g. from the Zurich SARS-CoV-2 Cohort study). Other parameters were only available on a national level, which may not reflect canton-specific differences adequately (e.g. process efficiency of CovidCode provision or MCT). In order to obtain more precise results (involving fewer assumptions), more granular and more regionally differentiated data from ongoing research studies are essential.

By evaluating the population at each step of the DPT notification cascade for the Canton of Zurich and in Switzerland, our analysis provides a first estimation of the contribution of SwissCovid to mitigating the pandemic. Our data suggest that the number of app notified persons receiving a quarantine recommendation corresponds to the equivalent of up to 5% of all mandatory quarantined contacts identified by MCT. Furthermore, we estimate that about 1 in 11 entered CovidCodes led to SARS-CoV-2 testing of an exposed proximity contact who was consecutively tested positive. Promoting app usage, increasing automation of the DPT notification cascade, and connecting rapid antigen testing results with the DPT notification cascade – while maintaining the privacy-preserving and voluntary nature of DPT – could further enhance the public health impact of the SwissCovid app.

## Data Availability

Non-public data are available from the corresponding author on reasonable request.

https://www.experimental.bfs.admin.ch/expstat/de/home/innovative-methoden/swisscovid-app-monitoring.html

## Acknowledgements

The authors wish to thank the Federal Office of Statistics and the Federal Office of Public Health for providing aggregated data.

